# Access to medicines for the treatment of chronic diseases in Chile: qualitative analysis of perceived patient barriers and facilitators in five regions of the country

**DOI:** 10.1101/2023.01.26.23285046

**Authors:** Carla Castillo Laborde, Isabel Matute, Ximena Sgombich, Daniel Jofré

**Author notes:** Corresponding Author Isabel Matute. These authors contributed equally to this work (Carla Castillo Laborde, Isabel Matute). These authors also contributed equally to this work (Ximena Sgombich, Daniel Jofré).

## Abstract

**Purpose:** To know patients’ perceptions of barriers and facilitators in access to medicines in general, and those associated with the treatment of three chronic conditions of high prevalence in Chile: diabetes, dyslipidemia and hypertension. Methods: Ten focus groups of patients with these diseases, diagnosed for at least six months and prescribed medication, were included. These were carried out in five regions of Chile: Arica, in the north, Aysén, in the south, and Valparaíso, Metropolitan, and Maule, in the central zone. The criteria for the formation of groups were region, age, health system (public or private) and socioeconomic level (residence commune). Sessions were recorded, transcribed, and analyzed by categories.

**Results:** The experience of access to medicines is determined by the insurance system, the experience of care with public or private providers, and geographical-administrative difference between capital and other regions. Beneficiaries of public sector, despite their greater socioeconomic vulnerability, perceive greater protection in access: access conditions, delivery reliability and adherence to pharmacological treatment are met. The main problem observed is the financing of treatments not covered by the health system. Policyholders in private sector perceive that they have access to medicines of better quality than those provided free of charge by public sector, but raise fears associated with the inability to afford them and distrust in the process, linked to alliances between laboratories and physicians. Public sector beneficiaries value territorial coverage of primary care, which guarantees access in isolated areas. Regarding the type of provider, public sector shows greater capacity for user loyalty, which is expressed in regular visits and follow-up; unlike discontinuous examinations among private sector beneficiaries.

**Conclusions:** Different access conditions both at the territorial level and in the health subsystems are evident. It is necessary to make progress in addressing the problem of access to medicines in a comprehensive manner.

## Introduction

Medications are one of the most widely used therapeutic tools in medicine; therefore, access is a challenge for health systems. Access to health services, and in this case to medicines, is understood as the opportunity for people to reach and obtain the appropriate services in situations of need (1), i.e., it implies a process that goes from the need for a medication, to its use by the patient (2). Access is a multidimensional concept, which includes availability, acceptability and geographic and financial accessibility as its most important dimensions (3,4). In other words, access to medicines implies their availability at a reasonable distance, their acceptance by patients and physicians and the real possibility of financing. These dimensions, as well as multiple barriers and facilitators associated with each of them, were also identified globally by a scope review on medications associated with three chronic conditions of high prevalence (5), finding that the most reported dimension, in countries of all regions and income levels, is affordability, and among the most relevant barriers, the availability of medications, long distances, cost, professional support and cultural aspects (5).

Despite the therapeutic importance of pharmaceutical drugs, the lack of access to essential medicines, defined as those that meet the priority health care needs of the population, is one of the most serious public health problems globally, affecting one third of the world’s population, with consequences for loss of life and deterioration of health (6).

In Chile, more than 57% of those over 15 years of age consume at least one medication, with paracetamol and acetyl salicylic acid being the most consumed nationwide, followed by losartan, metformin and atorvastatin (7). Of those with a health condition who receive medical care, 7.4% of people report having problems related to the delivery of medications to the health facility or access to them due to their cost (8). Medications represent the most important item of out-of-pocket spending on health (9); in fact, it is estimated that 30% of Chileans have stopped buying medications or stopped their treatments because they cannot afford them (10,11).

In the case of Chile, there is the public subsystem (FONASA beneficiaries), the private one (ISAPRE beneficiaries), and that of the Armed Forces, presenting a marked selection of risks, which leaves FONASA, for the most part, with the most vulnerable and needy population, and ISAPRE with the richest and healthiest population (12). There are vertical programs associated with certain diseases or conditions (13); that is, if we consider the conditions associated with the most consumed pharmaceutical drugs (losartan/hypertension, metformin/diabetes, atorvastatin/dyslipidemia), we can identify explicit guarantees (GES or AUGE) of access, opportunity, financial protection and quality for diabetes and hypertension, both for the beneficiaries of FONASA and those of ISAPRE (free delivery in the case of the former, and with a limited copayment established by law for the latter) (14), and we also identify a program called the Pharmacy Fund (FOFAR), for the FONASA beneficiaries, which ensures the availability (and free delivery) of medicines associated precisely with diabetes, hypertension and dyslipidemia in primary health care facilities (15). Thus, the way in which Chileans access their medicines depends, among others, on the subsystem to which they belong and the health condition from which they suffer.

In the context described above, the present study aimed to know the patients’ perceptions of the barriers and facilitators in access to medicines in general, and in particular regarding those associated with the treatment of three chronic conditions of high prevalence: diabetes, dyslipidemia and hypertension.

## Materials and Methods

### Participants and context

Ten focus groups were held between January and March 2021 in 5 regions of Chile: Arica and Parinacota (one) in the north, Aysén (one) in the south, and Valparaíso (one), Metropolitan (five) and Maule (two) in the central area.

The composition of the groups was defined according to: region, sex (both), age (45-64 and over 65 years), health care system (FONASA and ISAPRE) and socioeconomic level of the municipality of residence (% multidimensional poverty) (16). In addition, the following inclusion criteria were considered: people diagnosed with diabetes, dyslipidemia or hypertension, with prescribed pharmacological treatment, treated in public or private facilities.

The contact and enrollment process was carried out by a specialized team, identifying the potential participants, who met the profile required for each group, through community networks, delivering an invitation to participate in person, and subsequently re-contacting by phone to answer questions and confirm availability.

The meetings were held in hotel meeting rooms specifically hired for these purposes and in community centers. All focus groups were in person, so, in consideration of the COVID-19 Pandemic, demanding prevention protocols were observed.

The focus group participants did so voluntarily and accepted the conditions for carrying out the activity, which was formalized with the signing of an informed consent. This study is part of the FONIS Project SA19|0174, which was approved by the Bioethics Committee of the Faculty of Medicine of Clínica Alemana, Universidad del Desarrollo.

Each session was recorded and transcribed for further analysis.

### Data Analysis

To address the set objective, three dimensions of inquiry were considered:

1. General perception of access to health.
2. Access to medicines, from five dimensions of access (3–5):
  - Availability: related to the physical existence of medicines,
  - Geographical accessibility: geographical distance between medicines and location of users who need them,
  - Affordability: the relationship between the price of medicines and the patients’ ability to pay,
  - Acceptability: acceptance of medicines by the population or social groups and factors that increase or decrease the likelihood of their use; including users’ attitudes towards suppliers and medicines, and suppliers’ attitudes towards user characteristics,
  - Adaptation: the relationship between how resources are allocated to deliver medicines to users and their ability to adapt.
3. Access barriers and facilitators. Barriers understood as factors that hinder the target population from obtaining and making appropriate use of the medications it requires; and facilitators as factors that help the target population make use of them.

The thematic analysis considered an initial stage of familiarization with the data, from the reading of the transcriptions and initial notes. Subsequently, initial categories or codes were established, oriented to the three dimensions mentioned above. A search for topics or reduction of information was carried out based on structuring topics related to the problem investigated. Subsequently, the topics were reviewed, based on a process of re-coding and conceptual delimitation with the support of the Atlas Software.ti 9 (17). Finally, the topics were definitively identified, and a logic tree of topics and subtopics was created.

Additionally, empathy maps (18) were prepared to outline and characterize types of users in relation to access to medicines, based on the health care subsystem.

## Results

The presentation of the results is divided into three subsections: 1) Description of the participants; 2) General perceptions on access to health, and the barriers and facilitators depending on the dimensions of access (3–5); and 3) the differences between the perceptions of the FONASA and ISAPRE beneficiaries regarding access to medicines.

### Participant Characteristics

In total, for the 10 group interviews, 73 interviewees participated, mostly women (61.6%), FONASA beneficiaries (65.8%), with an average age of 60.2 years. In geographical terms, almost half of those interviewed belonged to the Metropolitan Region, followed by 22% in Maule (and rural area), and about 10% in the rest of the regions. Finally, 42.5% of the interviewees resided in communes with a high percentage of multidimensional poverty (Table 1).

**Table 1.**
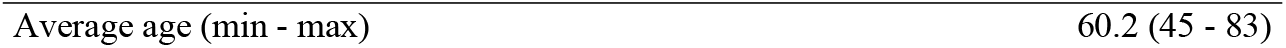
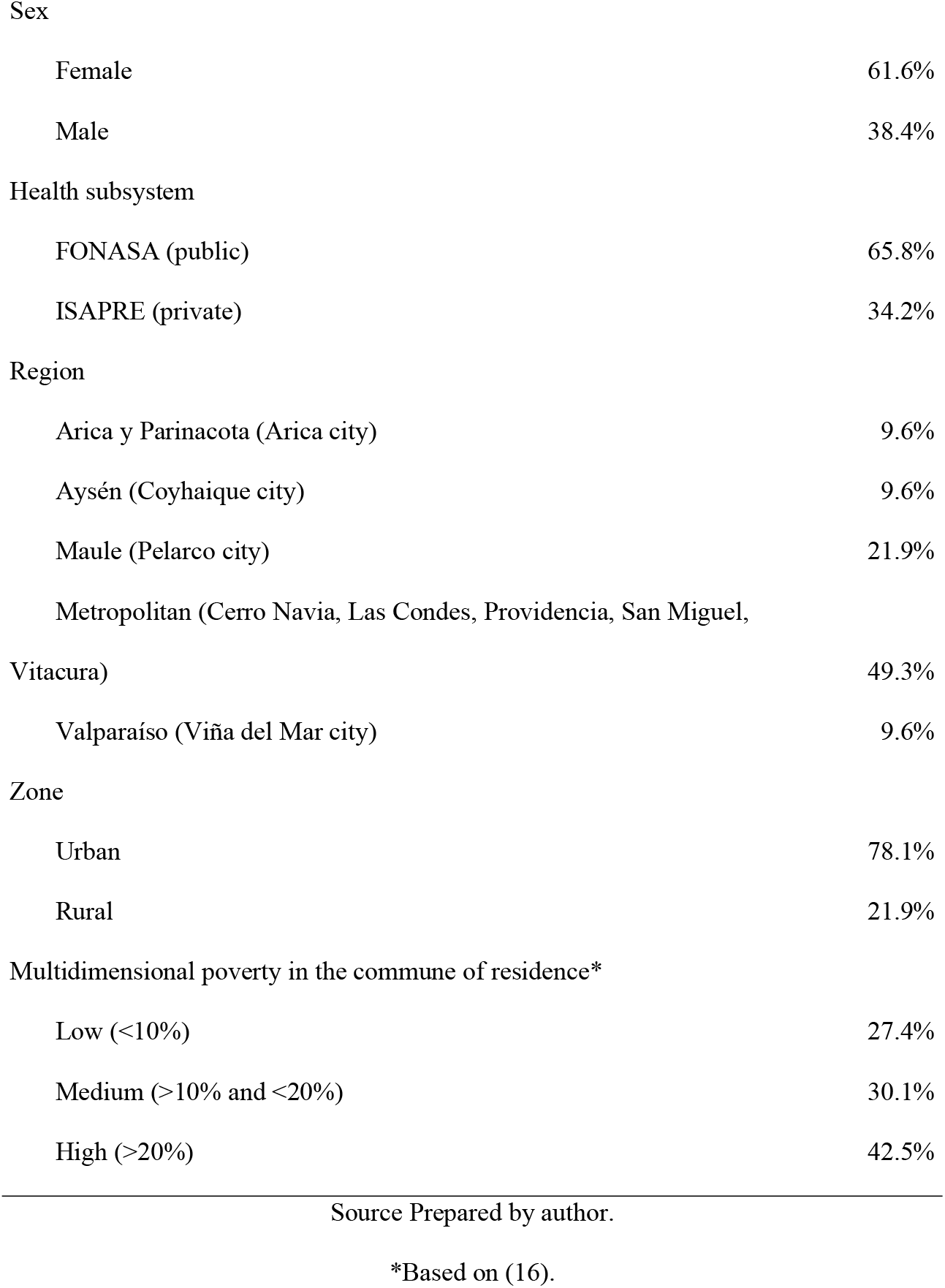
Participant Characteristics (n=73)

|  |  |
| --- | --- |
| Average age (min - max) | 60.2 (45 - 83) |

|  |  |  |
| --- | --- | --- |
| Sex |  |  |
|  | Female | 61.6% |
|  | Male | 38.4% |
| Health subsystem |  |  |
|  | FONASA (public) | 65.8% |
|  | ISAPRE (private) | 34.2% |
| Region |  |  |
|  | Arica y Parinacota (Arica city) | 9.6% |
|  | Aysén (Coyhaique city) | 9.6% |
|  | Maule (Pelarco city) | 21.9% |
|  | Metropolitan (Cerro Navia, Las Condes, Providencia, San Miguel, Vitacura) | 49.3% |
|  | Valparaíso (Viña del Mar city) | 9.6% |
| Zone |  |  |
|  | Urban | 78.1% |
|  | Rural | 21.9% |
| Multidimensional poverty in the commune of residence* |  |  |
|  | Low (<10%) | 27.4% |
|  | Medium (>10% and <20%) | 30.1% |
|  | High (>20%) | 42.5% |

Source Prepared by author.
\*Based on (16).

### Access dimensions

Below is a summary of the findings on the five access dimensions presented in the previous section (availability, geographical accessibility, affordability, acceptability, adaptation), within which, where possible, they are analyzed in terms of three cross-cutting axes identified as markers of the experience of access to medicines: the insurance system; ii) the experience of care with public and private providers; iii) and the geographical differences (Metropolitan Region/other regions; urban/rural) and other regions.

### Availability

In general terms, for the medicines associated with the three conditions of interest, no stock shortages are identified in the case of FONASA beneficiaries, although sometimes brief delays are observed.

> *“Moderator: Have you been told that the clinic pharmacy has no stock of medications?*
>
> *All: No*.
>
> *Woman (W): Every month we have a day*.
>
> *Moderator: So how difficult is it for you to access these medications? Man (M): It’s easy for me*.
>
> *W: No, it’s not difficult*.
>
> *Moderator: And has the current pandemic situation affected access to medicines?*
>
> *All: No*.
>
> *M: On the contrary, it has been become more streamlined*.*” (FG Arica, mixed, FONASA, 45-65 years old)*
>
> *“W: There are medicines that are sometimes missing, just have to wait*
>
> *W: Wait until they arrive, no longer than a week*.*” (FG Coyhaique, mixed, FONASA, 45 to 65 years old)*

However, the scenario changes when it comes to other health conditions that are more complex. In these cases, stock shortages are reported, rarely, which force people to buy in retail pharmacies.

> *“Moderator: So why do you sometimes have to buy them at the pharmacy and not at the clinic?*
>
> *W: When they do not arrive*.
>
> *W: For the problem I have in my spine, I must take an anti-inflammatory, and they don’t stock it, only Tramadol and Tramadol hurts me, because I have heart problems*.*” (FG Cerro Navia, mixed, FONASA, 45 to 65 years old)*

### Geographical accessibility

In the case of FONASA beneficiaries, the large extension and distribution throughout the national territory of the primary health care network, through which access is preferential, and which was reinforced by the delivery of medicines to homes during the pandemic, is noteworthy.

> *“Moderator: Who had their medications delivered to them during the pandemic? All: I did*.
>
> *Moderator: Don Juan, explain a little bit about that, before the pandemic, did they deliver medicines or not?*
>
> *M: No*.
>
> *M: Then, they began to deliver medications during the pandemic, and later in the clinic, they said it is a good idea for people over 65 years old*.
>
> *W: Until December that happened in my case. I had to go again in January*.*” (FG Coyhaique, mixed, FONASA, 45 to 65 years old)*

In the case of ISAPRE beneficiaries, access to medicines is preferably through retail pharmacies and, in recent years, through popular pharmacies (which emerged, in 2015, in the municipal context, and which have access to lower prices because they buy through the National Supply Center - CENABAST, like public sector facilities) (19).

> *“M: I buy in pharmacies, but also my wife, who is very much a “go-getter,” signed up in the Vitacura pharmacy, and there we buy it in the Vitabotica. We buy it at half price, less than half price. The truth is super convenient and the other*.
>
> *M: Which one?*
>
> *W: The Vitabotica*.
>
> *M: La Vitabotica”. (FG Vitacura – Las Condes, mixed, ISAPRE, 45 to 65 years old)*

People living in rural areas have some difficulties in accessing their medicines, especially due to periodicity, cost and travel time to clinics.

> *“M: Of course it is complicated for me, because in this time that we are with*… *last week that we were quarantined, many times I could not even come to church, because as I say, I am putting a lock on the gate, the bus and collective taxis pass by me, because they come full from Talca. I live in a rural area near her, but it’s different, because she has a car” (FG Pelarco, mixed, FONASA, over 65 years old)*

The older adults, meanwhile, express and demand the need for prioritized care in retail.

> *“M: From the beginning I wanted to make a clarification, there could be a difference in the pharmacies of the older adults, that is, when you go to the pharmacy today, there are 20 people in front of you, that is, it is easy to be 45 minutes in a line, and more, and I could, at least with an identity card, tell you, Hey, I’m an older adult and don’t make me wait 45 minutes*.*” (FG Providencia – Las Condes-San Miguel, mixed, ISAPRE, over 65 years old)*

### Affordability

In the case of FONASA beneficiaries, there is recognition and assessment of free access to medicines associated with the three conditions of interest.

> *“Moderator: And have any of you stopped taking medication because you couldn’t afford it?*
>
> *All: No*.
>
> *Moderator: And yours is accessible. How much do you spend on diabetes, hypertension and cholesterol triglyceride medications?*
>
> *M: I don’t spend anything W: Me neither*.
>
> *M: They give us all the medications*.
>
> *W: Sure, they give them. (FG Arica, mixed, FONASA, 45 to 65 years old)*
>
> *“W: Here the one who has money treats the disease he has, and we have to go here to the polyclinic that is the closest, and we do not have money to pay the expenses of the medicine we need, that’s why here in Chile it is bad, very bad “(FG Pelarco, mixed, FONASA, 45 to 65 years old)*

On the other hand, the FONASA user accesses a restricted set of medications (prioritized for coverage) that does not necessarily cover all their health problems. Those health problems whose medicines cannot be afforded are left unattended or partially resolved. This situation is rare when it comes to hypertension, dyslipidemia and diabetes, but it is observed in the management of side effects and in the management of other more complex diseases.

> *W: But later I started to feel well without a problem, but I had to stop buying them if I couldn’t afford it, a box with 12 pills cost me $ 57,000. Then I could not do it, it was day-to-day that I continued with the polyclinic, but they lowered the dose and healed me of my ulcers, and I continue with my medicine from the polyclinic, I will have to continue because I do not have the resources to buy a medication of 50 and so thousand every month. “ (FG Coyhaique, mixed, FONASA, 45 to 65 years old)*
>
> *“W: I think so, because, because, for example, here in the clinic they tell you Metformin and you get Metformin, but if you go to the market, to the pharmacies, it is called Hipoglucin, and that medication is so coated that it does not produce the effects that Metformin produces*.
>
> *W: Yes*.
>
> *W: Because of the stomach*.*” (FG Cerro Navia, mixed, FONASA, 45 to 65 years old)*

The GES/AUGE corresponds to a policy that, among others, allows to reduce the economic burden linked to the treatment of diseases such as diabetes and hypertension. Paradoxically, people do not always associate free medicines with this public policy, that is, their direct consequences (availability of medicines) are known but their origin in the GES (or in FOFAR, in the case of dyslipidemia) is not necessarily identified. In practice, this policy is so relevant that the most economically vulnerable groups could not attempt not to have free medicines, at least for hypertension, diabetes and dyslipidemia.

> *“Moderator: Do any of you receive these medicines from GES?*
>
> *W: No, what is that?*
>
> *Moderator: Do any of you receive medication through GES? Several ask: What is it?*
>
> *W: what is GES?” (FG Coyhaique, mixed, FONASA, 45 to 65 years old)*

In the case of ISAPRE beneficiaries, in general they pay directly for medications for hypertension, diabetes and dyslipidemia, predominating in this group the idea that free access refers to socially vulnerable groups and cared for in the primary health care network, clearly appearing the concept of social class different from their own: “That class is the most vulnerable”.

> *“M: I believe that the vast majority buy them. W: You just have to buy them*.
>
> *M: Out of your pocket*.
>
> *M: I think there is a very small class, and that class is the one that is most vulnerable, it is the one that resorts to primary health care which are the SAMU, or SAPU, I do not know what they are called now*.
>
> *M: But most I think still resort to buying them with their money, in a particular way*.*” (FG Viña del Mar, mixed, ISAPRE, 45 to 65 years old)*

For this group of beneficiaries, there is a lack of knowledge and low use of GES/AUGE and the medication coverage it offers; they preferentially choose to maintain freedom of purchase and their relationship with the treating doctors. In ISAPRE the GES/AUGE are attended by a closed network of private providers, defined a priori by each insurer, therefore, they restrict the free choice or continuity of treatment with the treating physician, before having to adapt to the GES protocols (or AUGE). The cost of medicines would be considered marginal relative to meeting the requirements of the plan enrollment. From the perception and beliefs of these users, there is a lack of information and understanding about the process of accessing GES, even some assume that they would have to pertain to the public health care system, which would be a barrier to access for these segments.

> *“Moderator: Have you made use of GES for these pathologies [diabetes, hypertension]?*
>
> *W: No*.
>
> *W: No*.
>
> *W: It is that my apprehension was ultimately, according to what I understand, they force you to a doctor from the Auge*…
>
> *W: In the ISAPRE*.
>
> *W: Sure, and ultimately if you want to go to your doctor, you can’t. M: Unless it just coincides*…
>
> *W: Ah, but that’s very difficult*.
>
> *W: But what you can do, for example, is go to the doctor from the Auge, to examine you, to give you the medicines, which are quite cheap, and then you tell your doctor, every so often, you know that they are giving me so much, go check*.
>
> *W: Ah, there you have a parallel*.
>
> *W: Sure*.*” (FG Vitacura – Las Condes, mixed, ISAPRE, 45 to 65 years old)*

In this way, the use of GES, for this group, would be justified in the case of pathologies with higher cost treatments.

> *“M: For the medication I take for the prostate issue, which is another issue, I do it through GES*.
>
> *W: Oh too*.
>
> *W: Ah, but you pay nothing there*.
>
> *M: Of course, it is a daily pill, which are 30. They cost $45,000. Moderator: $45*.*000?*
>
> *M: And if not a little more. And on the other hand, five. W: Wow, I mean, I do all the management anyway*.
>
> *W: You pay $5,000 monthly*.
>
> *W: What happens is that your pathology was in the GES, in the Auge, so you register, that the doctor registers you for the Auge, you call the ISAPRE and the ISAPRE refers you to a doctor from the Auge and you must pay that monthly $ 5,000*.*” (FG Vitacura*
>
> *– Las Condes, mixed, ISAPRE, 45 to 65 years old)*

On the other hand, it is recognized, among this group of users, that GES operates as a conditional transfer, that is, the delivery of medicines requires the person to go through systematic health checkups.

> *“M: Of course, I go every 6 months to the doctor, they examine me, they still do exams, they force you to be monitoring, which is a bit the idea, because ultimately people let themselves go a bit*.*” (FG Vitacura – Las Condes, mixed, ISAPRE, 45 to 65 years old)*
>
> Another element that interferes with people’s access to GES coverage is the role played by doctors, who can act as facilitators or hinder access to GES.
>
> *“W: They do not tell you, there are doctors who do and others who do not, for example, I always went to the ophthalmologist and, well, I was going for the driver’s license that previously took away a year for not wearing glasses, but pretentiously, not anymore, so I went to the ophthalmologist, well she happily* … *I, exceptional, because she told me “you know what, you can come in one more week”, Yes, I told her, but why “because in five more days you turn 65 and I can give them to you through the GES”, but because she proposed it to me, because otherwise I would have had no idea”. (FG Providencia -Las Condes – San Miguel, mixed, ISAPRE, over 65 years old)*

In general terms, beyond the differences in terms of the health subsystem to which the beneficiaries belong, and the three chronic conditions of interest (whose medicines they access either free or at low cost), in terms of affordability, the existence of a precarious balance in terms of access to medicines is recognized among the participants. The occurrence of a simple event, such as the emergence of diseases that affect other members of the family group, or other diseases with treatments not covered or of higher cost, is enough to destabilize the family budget, and often force them to prioritize.

> *W: One has a relatively easy time, but it is enough that someone gets sick, I do not know, an illness, my daughter needed antibiotics and 50 thousand pesos an antibiotic, more, I do not know, other things and the budget, apart from what I am taking fixed every month, there is no budget that endures in terms of medicines. Any family can be economically destabilized in a month with an illness, in a month. One can overcome it, but it is in installments, you get indebted to get out of that and that’s how it is*.*” (FG Providencia – Las Condes, mixed, FONASA and ISAPRE, 45-65 years old)*
>
> Finally, from the point of view of affordability, a positive assessment towards Popular Pharmacies is also perceived.
>
> *W: I will always buy a medicine that is for osteoarthritis that in the pharmacy costs about $ 27,000 and here it costs 16, it is a tremendous, what is it called, savings, and there are many people who do not know that type of help*.
>
> *W: That’s super important and valuable because you save a lot of money that you can spend on food or milk, on anything, so*…
>
> *W: It has to say permanent prescription and has nothing to do with social class, only medication and diagnosis. “ (FG Coyhaique, mixed, FONASA, 45 to 65 years old)*

### Acceptability

FONASA beneficiaries, who receive free medicines through the network of public providers, are confident that these will yield benefits for their health, and in some cases the quality is even considered to have improved over time.

> *“Moderator: So, about the quality and effectiveness of the medications you receive in the clinic. How do you evaluate them? What do you think of the medications you are given?*
>
> *M: To me they are good. W: Same*.
>
> *W: Very good too*.
>
> *M: Good because the doctor, although it is true, gives you the prescription and everything. It’s the exact thing, what’s needed, the corresponding doses, everything. No problem*.
>
> *M: Yes, very good*.
>
> *Moderator: And do you think the medications from the clinic could do any harm? Or not?*
>
> *M: No*
>
> *Moderator: Do you trust your medications 100%?*
>
> *M: Yes, because it has changed a lot. Before there were quite a few Chilean laboratories, but much has changed. The medications have changed*.*” (FG Arica, mixed, FONASA, 45 to 65 years old)*

However, although the effectiveness of medicines for the diseases studied is not questioned, in the case of Metformin the side effects are suffered, which result in additional health expenses to overcome them. This situation is reported in several groups of users served in the public health care network.

> *“W: I already told you, they did X-rays, ultrasound scans, the whole story. What happened: consequently, I got ulcers with metformin, apart from the fact that it is a very large pill, it is very unpleasant to take it, and you must break it and the whole story. Then the doctor told me, he gave me the alternative, unfortunately I cannot give you another medication because we are a polyclinic*.
>
> *W: Now, I started to take Metformin again, and it makes me vomit and gives me diarrhea*.*” (FG Coyhaique, mixed, FONASA, 45 to 65 years old)*

On the other hand, among the ISAPRE beneficiaries, higher prices are associated with greater effectiveness, perceiving a higher quality of the medicines they buy in retail pharmacies, than those received by FONASA beneficiaries in the public sector. In this sense, belonging to an ISAPRE would be a guarantee of access to medicines that are considered more expensive, but also more effective.

> *“Moderator: Do you think that having ISAPRE or FONASA changes access to medicines?*
>
> *W: Yes*.
>
> *Moderator: Why, how?*
>
> *W: Because the doctor, in fact, you see, can give him or herself a greater luxury of giving you better qualities medications, of not being in an ISAPRE because, for example, you go to the clinic, and they give you the cheapest medications there are*…
>
> *W: The generics*.
>
> *W: That is, generics, which supposedly they say are not the same quality as medications that have a name or a pharmacy, that is, from a better laboratory, I do not know, it has always been said. “ (FG Providencia – Las Condes – San Miguel, ISAPRE, over 65 years old)*

Finally, ISAPRE users express greater reluctance to use bioequivalent medicines, with suspicion predominating in their discourse about the effectiveness and quality of generic and bioequivalent medications. This articulates some beliefs and opinions based on own experiences of use: that they work like placebos, that doses must be increased to reach the effectiveness of a brand-name drug, that the savings may be less if doses are increased.

> *“W: They are not the same either, one can notice it even in terms of sensations, in the basics, paracetamol for headache, you have the generic, the bioequivalent and the name brand and with the last two your headache goes away and the other, you have to take 3 or 4 pills and in the end what is the generic for if it does not work for you, if it doesn’t give you the result. You say, I spent two more coins or two less coins, but in the end, as I took more, the generic came out more expensive*.*” (FG Providencia – Las Condes, mixed, FONASA and ISAPRE, 45-65 years old)*

### Adaptation

In general, people served in the public network evaluate that the dynamics of medication provision is in line with their possibilities. Medicine pickup is a regular activity that people carry out every month and that keeps them in contact with the health care system. The extensive territorial network of health facilities addresses the issue of physical distance being accessible to users.

> *“Moderator: And how often, how many times*… *How often do you actually go to the clinic to get your medication?*
>
> *W: Every month. M: Each month. W: Every month*.
>
> *W: Every month we have a day*.
>
> *Moderator: So how difficult is it for you to access these medications? M: It’s easy for me*.
>
> *W: No, it’s not difficult*.*” (FG Arica, mixed, FONASA, 45 to 65 years old)*

The logistics of medicine provision is one of the most recognized aspects of primary care; there are significant improvements in terms of access and treatment, and it introduces a differentiation between public clinics and hospitals, where the latter represent the most critical face of the public health system.

The adaptation is lower in very particular cases that are referred to groups, such as people who live alone and have lost autonomy, older adults with disabilities or people living in isolated places, these segments can evaluate their access possibilities as restricted.

> *“M: But in general, the great majority of people are very few who are isolated, very few, the great majority have good services*.*” (FG Coyhaique, mixed, FONASA, 45 to 65 years old)*
>
> *“W: One has to remember to go to pick up the medications, if not, he or she loses them, that’s where I tell him about the elderly, let’s say, there are old people who are not*… *they are old, and they cannot come to pick them up, they have no young person to send” (FG Pelarco, FONASA, 45 to 65 years old)*

### FONASA and ISAPRE differences

Figures 1 and 2 present the empathy maps associated with the FONASA and ISAPRE beneficiaries, respectively.

**Figure 1.**
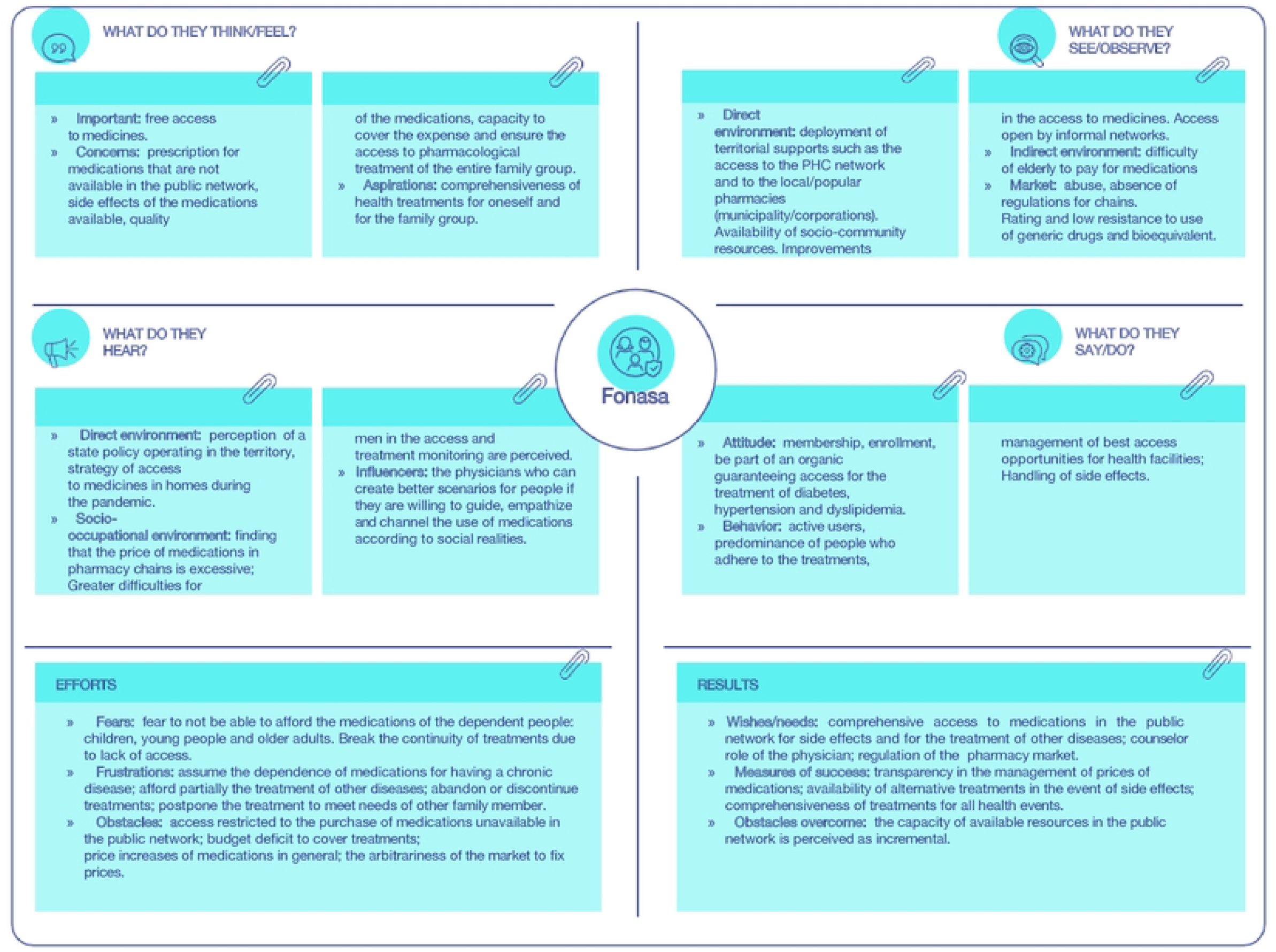
FONASA Beneficiary Empathy Map.

**Figure 2.**
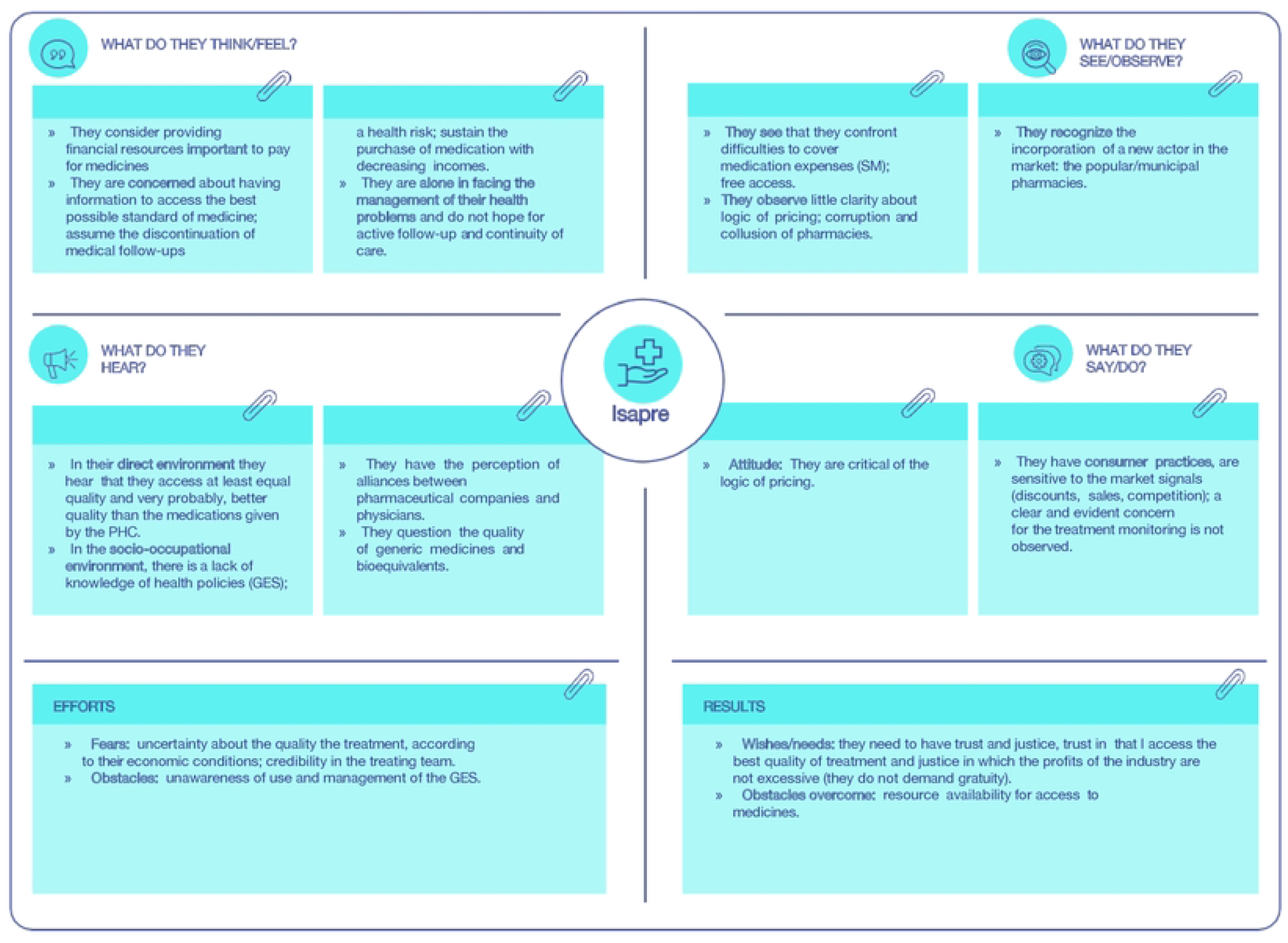
ISAPRE Beneficiary Empathy Map.

According to that presented in Figure 1, it is possible to note that FONASA beneficiaries, despite their greater socio-economic vulnerability, perceive greater protection in access to their medicines. Access conditions, delivery regularity and adherence to pharmacological treatment are met. For this group, the treatment financing for comorbidities and morbidities of the family group is perceived as the main problem.

On the other hand, users in the public sector value the territorial coverage of primary care, which guarantees access in isolated areas. Regarding the type of provider, the public sector shows greater capacity for loyalty of users, which is expressed in regular check-ups and feeling of monitoring; unlike more irregular visits among beneficiaries of the private sector.

As observed in Figure 2, private sector policyholders, for their part, understand that they have access to better quality medicines than those provided free of charge by the public sector. However, they report fears associated with inability to afford them and greater distrust in the process, linked to alliances between laboratories and physicians.

## Discussion

The findings of the present study characterize the access to medicines in Chile of people of different socioeconomic conditions and residents in scattered regions of the country, for the three chronic conditions of interest: hypertension, diabetes and dyslipidemia. As mentioned in the introductory section, these three non-communicable conditions have a high prevalence in Chile and, from the point of view of coverage by the health system, they have been prioritized, through the Explicit Health Guarantees – GES, for the FONASA and ISAPRE beneficiaries, and the Primary Health Care Pharmacy Fund, for the FONASA beneficiaries (20).

The high segmentation of the Chilean health system, the geographical extension of the country, as well as the existence of rural and isolated areas throughout the territory significantly determine the way Chileans access their medicines, as well as the barriers they must face, and the facilitators they have in the process.

The provision of medicines is affected by variables in the organization of health services. It is noteworthy that primary care has succeeded in establishing coordination and monitoring of the population caring for, at least for chronic diseases, which would favor the control of these diseases.

The above explains that, in general terms, people affiliated with FONASA have access to medicines for the treatment of the diagnoses studied through the institutional public health network, free of charge because of public policies that include: Explicit Health Guarantees (GES or AUGE) and Pharmacy Fund (FOFAR). For their part, the people affiliated with ISAPRE mostly access medications through direct purchase in pharmacies and pay for the entire treatment, which is considered an insignificant expense due to the low cost of these in the case of the three conditions studied. This segment can also access the GES; however, a marginal use of guarantees was observed associated with administrative barriers, ignorance and resistance to attend the preferential network of ISAPRE for the GES, which implies changing the treating physician.

In general terms, there are no relevant barriers in the case of medicines for the three conditions of interest for FONASA beneficiaries. However, access barriers for other medications are identified and these are associated, in the case of public sector beneficiaries, with out-of-pocket spending on medicines not covered or not prioritized by the health system and, in those cases where the medicines are covered but circumstantially encounter stock shortages in public establishments, the availability barrier appears, with the consequent affordability problems associated with having to purchase them in retail. The facilitators, for their part, include policies and programs that establish medication coverage and guarantee availability and financial protection; the territorial deployment of primary care in the country; and the acceptance and assessment by FONASA users of the public sector care model, which determines the control and permanent contact of users with the health system.

In the case of private sector beneficiaries who, as mentioned, buy mainly in retail, significantly associating the price of quality medications; one of the barriers identified is adaptation, and has to do with the non-acceptance or rejection by users of retail practices (lack of transparency, collusion, etc.), which in turn is related to the out-of-pocket expenses they must incur. In the case of these beneficiaries, another adaptation barrier also emerges, associated with the lack of knowledge of the processes for obtaining the benefits delivered by the GES, and another of acceptability, related to the rejection of the conditions imposed for obtaining them (e.g., bureaucracy, renunciation of free choice, use of generics). Among the facilitators for the ISAPRE users, there is the budget availability that, in general, allows them to pay for the required medicines, at least for the three chronic conditions of interest for the analysis, the potential lower out-of-pocket expenditure to be able to access through the GES, and the possibility of having complementary coverage through voluntary insurance.

It is worth mentioning that the eruption of popular or municipal pharmacies is an access facilitator recognized by users in a transversal way, allowing access to lower prices compared to retail.

The results presented here are in line with findings published in the literature, first, regarding the relevance, as a facilitator for access to medicines, of having coverage or health insurance, from the point of view of out-of-pocket spending and affordability, and as a barrier if this possibility is not available. It is possible to find experiences of countries in different economic or geographical contexts in this regard (21–24). In fact, in the Americas region, there are close examples, such as the case of Brazil with the free delivery of medications by the SUS, and its Health Has no Price programs (for hypertension and diabetes as is of interest in this research) and Popular Pharmacies (25–28). Another barrier that emerges in the study and is also reported in the literature for various countries, such as Barbados, the US and Colombia, relates to the bureaucracy or how complex and cumbersome it is to understand the insurance requirements to obtain medication coverage (29–31), as happens in Chile to the ISAPRE beneficiaries for the use of GES.

One of the main limitations of this study is related to the conduct of group interviews during the Covid-19 pandemic, which, among others, required strict monitoring of protocols, and also made it necessary to address, within the framework of the interviews, some issues related to access to medication during the pandemic, and then further examine (removing the focus of the pandemic) more general issues associated with this access. However, this limitation became, in turn, an opportunity to explore people’s perceptions about access to medicines in the pandemic context, and the ways in which the Chilean health system addressed this problem in this period of greatest difficulties.

Finally, within the framework of this research it was possible to identify opportunities for improvement for public health policy, among the main ones, and in line with the aforementioned, is the importance of moving towards comprehensive medicine coverage, as well as its free delivery by health systems to achieve better and more equitable access. Another implication relates to the need to strengthen, on the part of the health authority, the messages around the quality of bioequivalents, to increase the confidence of users in the policies of access to generics, which are delivered free of charge by the system. Finally, it is necessary to inform the population about how to access the benefits of policies or programs, and to make the relationship between gratuity and these policies transparent, as part of the State’s commitment to advancing the rights of access to health.

## Data Availability

Once accepted, transcripts from the focus groups will be uploaded to a public repository and the URL will be shared with PLOS ONE. Transcripts are in Spanish only.

## Acknowledgements

The authors thank the financing of the FONIS SA19|0174 Project, granted by the National Agency for Research and Development, Chile (ANID), in the framework of which this study was carried out.

The authors also thank the fieldwork team for their support in conducting the interviews.

Finally, we thank all the participants in the interviews conducted for sharing their time and experiences.

